## Appendix 1 Search Strategy for "Sexual and Reproductive Health Needs of Women with Severe Mental Illness in Low- and Middle-Income Countries: A Scoping Review"

MEDLINE

1150 found – duplicates removed 1148

<https://ovidsp.ovid.com/ovidweb.cgi?T=JS&NEWS=N&PAGE=main&SHAREDSEARCHID=2tJuhTDWafChmLcrAUiP75g631QskcZj1NJk8oja1ZIKSRV8CnzwVOAQnJxbAGMbD>

1

sexual health.mp. or Sexual Health/ or HIV Infections/ or Sexually Transmitted Diseases/ or Sexual Behavior/

00:02

2

Family Planning Services/ or reproductive health.mp. or Reproductive Health/ or Abortion, Induced/ or Reproductive Health Services/

00:01

3

sexual violence.mp. or Sex Offenses/

00:01

4

Hormonal Contraception/ or Contraception, Postcoital/ or contraception.mp. or Contraception Behavior/ or Long-Acting Reversible Contraception/ or Contraception, Barrier/ or Contraception/ or Contraception, Immunologic/

00:01

5

Sexual Behavior/ or sexual identity.mp.

00:01

6

Sexual Behavior/ or sexual dysfunction.mp. or Sexual Dysfunctions, Psychological/

00:01

7

HIV/ or hiv.mp.

00:01

8

Bipolar Disorder/ or Psychotic Disorders/ or Mental Disorders/ or severe mental illness.mp. or Schizophrenia/

00:01

9

Developing Countries/

00:01

10

(afghanistan or albania or algeria or american samoa or angola or "antigua and barbuda" or antigua or barbuda or argentina or armenia or armenian or aruba or azerbaijan or bahrain or bangladesh or barbados or republic of belarus or belarus or byelarus or belorussia or byelorussian or belize or british honduras or benin or dahomey or bhutan or bolivia or "bosnia and herzegovina" or bosnia or herzegovina or botswana or bechuanaland or brazil or brasil or bulgaria or burkina faso or burkina fasso or upper volta or burundi or urundi or cabo verde or cape verde or cambodia or kampuchea or khmer republic or cameroon or cameron or cameroun or central african republic or ubangi shari or chad or chile or china or colombia or comoros or comoro islands or iles comores or mayotte or democratic republic of the congo or democratic republic congo or congo or zaire or costa rica or "cote d’ivoire" or "cote d’ ivoire" or cote divoire or cote d ivoire or ivory coast or croatia or cuba or cyprus or czech republic or czechoslovakia or djibouti or french somaliland or dominica or dominican republic or ecuador or egypt or united arab republic or el salvador or equatorial guinea or spanish guinea or eritrea or estonia or eswatini or swaziland or ethiopia or fiji or gabon or gabonese republic or gambia or "georgia (republic)" or georgian or ghana or gold coast or gibraltar or greece or grenada or guam or guatemala or guinea or guinea bissau or guyana or british guiana or haiti or hispaniola or honduras or hungary or india or indonesia or timor or iran or iraq or isle of man or jamaica or jordan or kazakhstan or kazakh or kenya or "democratic people’s republic of korea" or republic of korea or north korea or south korea or korea or kosovo or kyrgyzstan or kirghizia or kirgizstan or kyrgyz republic or kirghiz or laos or lao pdr or "lao people's democratic republic" or latvia or lebanon or lebanese republic or lesotho or basutoland or liberia or libya or libyan arab jamahiriya or lithuania or macau or macao or republic of north macedonia or macedonia or madagascar or malagasy republic or malawi or nyasaland or malaysia or malay federation or malaya federation or maldives or indian ocean islands or indian ocean or mali or malta or micronesia or federated states of micronesia or kiribati or marshall islands or nauru or northern mariana islands or palau or tuvalu or mauritania or mauritius or mexico or moldova or moldovian or mongolia or montenegro or morocco or ifni or mozambique or portuguese east africa or myanmar or burma or namibia or nepal or netherlands antilles or nicaragua or niger or nigeria or oman or muscat or pakistan or panama or papua new guinea or new guinea or paraguay or peru or philippines or philipines or phillipines or phillippines or poland or "polish people's republic" or portugal or portuguese republic or puerto rico or romania or russia or russian federation or ussr or soviet union or union of soviet socialist republics or rwanda or ruanda or samoa or pacific islands or polynesia or samoan islands or navigator island or navigator islands or "sao tome and principe" or saudi arabia or senegal or serbia or seychelles or sierra leone or slovakia or slovak republic or slovenia or melanesia or solomon island or solomon islands or norfolk island or norfolk islands or somalia or south africa or south sudan or sri lanka or ceylon or "saint kitts and nevis" or "st. kitts and nevis" or saint lucia or "st. lucia" or "saint vincent and the grenadines" or saint vincent or "st. vincent" or grenadines or sudan or suriname or surinam or dutch guiana or netherlands guiana or syria or syrian arab republic or tajikistan or tadjikistan or tadzhikistan or tadzhik or tanzania or tanganyika or thailand or siam or timor leste or east timor or togo or togolese republic or tonga or "trinidad and tobago" or trinidad or tobago or tunisia or turkey or turkmenistan or turkmen or uganda or ukraine or uruguay or uzbekistan or uzbek or vanuatu or new hebrides or venezuela or vietnam or viet nam or middle east or west bank or gaza or palestine or yemen or yugoslavia or zambia or zimbabwe or northern rhodesia or global south or africa south of the sahara or sub-saharan africa or subsaharan africa or africa, central or central africa or africa, northern or north africa or northern africa or magreb or maghrib or sahara or africa, southern or southern africa or africa, eastern or east africa or eastern africa or africa, western or west africa or western africa or west indies or indian ocean islands or caribbean or central america or latin america or "south and central america" or south america or asia, central or central asia or asia, northern or north asia or northern asia or asia, southeastern or southeastern asia or south eastern asia or southeast asia or south east asia or asia, western or western asia or europe, eastern or east europe or eastern europe or developing country or developing countries or developing nation? or developing population? or developing world or less developed countr* or less developed nation? or less developed population? or less developed world or lesser developed countr* or lesser developed nation? or lesser developed population? or lesser developed world or under developed countr* or under developed nation? or under developed population? or under developed world or underdeveloped countr* or underdeveloped nation? or underdeveloped population? or underdeveloped world or middle income countr* or middle income nation? or middle income population? or low income countr* or low income nation? or low income population? or lower income countr* or lower income nation? or lower income population? or underserved countr* or underserved nation? or underserved population? or underserved world or under served countr* or under served nation? or under served population? or under served world or deprived countr* or deprived nation? or deprived population? or deprived world or poor countr* or poor nation? or poor population? or poor world or poorer countr* or poorer nation? or poorer population? or poorer world or developing econom* or less developed econom* or lesser developed econom* or under developed econom* or underdeveloped econom* or middle income econom* or low income econom* or lower income econom* or low gdp or low gnp or low gross domestic or low gross national or lower gdp or lower gnp or lower gross domestic or lower gross national or lmic or lmics or third world or lami countr* or transitional countr* or emerging economies or emerging nation?).ti,ab,sh,kf.

11

9 or 10

12

Maternal Health Services/ or Pregnancy/ or Depression, Postpartum/ or motherhood.mp. or Mothers/

13

1 or 2 or 3 or 4 or 5 or 6 or 7 or 12

14

8 and 11 and 13

EMBASE

(terms as per MEDLINE) 960 – duplicates removed 951

CINAHL

“sexual health” OR HIV OR “sexually transmitted disease” OR “sexual behaviour” OR “family planning” OR “reproductive health” OR abortion OR “sexual violence” OR contraception OR “sexual identity” OR “sexual dysfunction” OR “pregnancy” OR “motherhood”

AND

“Severe mental illness” OR schizophrenia OR bipolar OR psychosis

AND

MH "Developing Countries" or MH "low and middle income countries" or MH "Afghanistan" or MH "Albania" or MH "Algeria" or MH "Angola" or MH "Argentina" or MH "Armenia" or MH "Azerbaijan" or MH "Bangladesh" or MH "Belarus" or MH "Belize" or MH "Benin" or MH "Bhutan" or MH "Bolivia" or MH "Bosnia-Herzegovina" or MH "Botswana" or MH "Brazil" or MH "Burkina Faso" or MH "Burundi" or MH "Cambodia" or MH "Cameroon" or MH "Cape Verde" or MH "Central African Republic" or MH "Chad" or MH "China" or MH "Colombia" or MH "Comoros" or MH "Congo" or MH "Costa Rica" or MH "Cote d'Ivoire" or MH "Cuba" or MH "Democratic Republic of Congo" or MH "Djibouti" or MH "Dominica" or MH "Dominican Republic" or MH "East Timor" or MH "Ecuador" or MH "Egypt" or MH "El Salvador" or MH "Equatorial Guinea" or MH "Eritrea" or MH "Ethiopia" or MH "Fiji" or MH "Gabon" or MH "Gambia" or MH "Georgia (Republic)" or MH "Ghana" or MH "Guatemala" or "MH "Guinea" or MH "Guinea-Bissau" or MH "Guyana" or MH "Haiti" or MH "Honduras" or MH "India" or MH "Indian Ocean Islands" or MH "Indonesia" or MH "Iran" or MH "Iraq" or MH "Jamaica" or MH "Jordan" or MH "Kazakhstan" or MH "Kenya" or MH "Kyrgyzstan" or MH "Laos" or MH "Lebanon" or MH "Lesotho" or MH "Liberia" or MH "Libya" or MH "Macedonia (Republic)" or MH "Madagascar" or MH "Malawi" or MH "Malaysia" or MH "Mali" or MH "Mauritania" or MH "Mexico" or MH "Micronesia" or MH "Moldova" or MH "Mongolia" or MH "Morocco" or MH "Mozambique" or MH "Myanmar" or MH "Namibia" or MH "Nepal" or MH "Nicaragua" or MH "Niger" or MH "Nigeria" or MH "North Korea" or MH "Pakistan" or MH "Panama" or MH "Papua New Guinea" or MH "Paraguay" or MH "Peru" or MH "Philippines" or MH "Polynesia" or MH "Rwanda" or MH "Samoa" or MH "Senegal" or MH "Serbia" or MH "Sierra Leone" or MH "Somalia" or MH "South Africa" or MH "Sri Lanka" or MH "Sudan" or MH Suriname or MH "Swaziland" or MH "Syria" or MH "Tajikistan" or MH "Tanzania" or MH "Thailand" or MH "Togo" or MH "Tunisia" or MH "Turkey" or MH "Turkmenistan" or MH "Uganda" or MH "Ukraine" or MH "Uzbekistan" or MH "Venezuela" or MH "Vietnam" or MH "Yemen" or MH "Zambia" or MH "Zimbabwe"

TI ("cabo verde*" or "democratic people's republic of korea" or eswatini or kiribati* or "gaza strip" or grenada or grenadian* or grenadines or kosov* or "lao people's democratic republic" or maldiv* or "marshall island*" or mauriti* or montenegr* or montserrat* or nauru* or niue* or palestin* or "saint helena*" or "saint lucia*" or "saint vincent*" or "sao tome*" or "solomon island*" or "south sudan*" or timorese or tokelau* or tonga* or tuvalu* or vanuatu* or "wallis and futuna" or "west bank") OR AB ("cabo verde*" or "democratic people's republic of korea" or eswatini or kiribati* or "gaza strip" or grenada or grenadian* or grenadines or kosov* or "lao people's democratic republic" or maldiv* or "marshall island*" or mauriti* or montenegr* or montserrat* or nauru* or niue* or palestin* or "saint helena*" or "saint lucia*" or "saint vincent*" or "sao tome*" or "solomon island*" or "south sudan*" or timorese or tokelau* or tonga* or tuvalu* or vanuatu* or "wallis and futuna" or "west bank")

46 results- duplicates removed 43

https://search-ebscohost-com.nottingham.idm.oclc.org/login.aspx?direct=true&db=rzh&bquery=(((TI+(%26quot%3bcabo+verde*%26quot%3b+OR+%26quot%3bdemocratic+people%26%2339%3bs+republic+of+korea%26quot%3b+OR+eswatini+OR+kiribati*+OR+%26quot%3bgaza+strip%26quot%3b+OR+grenada+OR+grenadian*+OR+grenadines+OR+kosov*+OR+%26quot%3blao+people%26%2339%3bs+democratic+republic%26quot%3b+OR+maldiv*+OR+%26quot%3bmarshall+island*%26quot%3b+OR+mauriti*+OR+montenegr*+OR+montserrat*+OR+nauru*+OR+niue*+OR+palestin*+OR+%26quot%3bsaint+helena*%26quot%3b+OR+%26quot%3bsaint+lucia*%26quot%3b+OR+%26quot%3bsaint+vincent*%26quot%3b+OR+%26quot%3bsao+tome*%26quot%3b+OR+%26quot%3bsolomon+island*%26quot%3b+OR+%26quot%3bsouth+sudan*%26quot%3b+OR+timorese+OR+tokelau*+OR+tonga*+OR+tuvalu*+OR+vanuatu*+OR+%26quot%3bwallis+and+futuna%26quot%3b+OR+%26quot%3bwest+bank%26quot%3b)+OR+AB+(%26quot%3bcabo+verde*%26quot%3b+OR+%26quot%3bdemocratic+people%26%2339%3bs+republic+of+korea%26quot%3b+OR+eswatini+OR+kiribati*+OR+%26quot%3bgaza+strip%26quot%3b+OR+grenada+OR+grenadian*+OR+grenadines+OR+kosov*+OR+%26quot%3blao+people%26%2339%3bs+democratic+republic%26quot%3b+OR+maldiv*+OR+%26quot%3bmarshall+island*%26quot%3b+OR+mauriti*+OR+montenegr*+OR+montserrat*+OR+nauru*+OR+niue*+OR+palestin*+OR+%26quot%3bsaint+helena*%26quot%3b+OR+%26quot%3bsaint+lucia*%26quot%3b+OR+%26quot%3bsaint+vincent*%26quot%3b+OR+%26quot%3bsao+tome*%26quot%3b+OR+%26quot%3bsolomon+island*%26quot%3b+OR+%26quot%3bsouth+sudan*%26quot%3b+OR+timorese+OR+tokelau*+OR+tonga*+OR+tuvalu*+OR+vanuatu*+OR+%26quot%3bwallis+and+futuna%26quot%3b+OR+%26quot%3bwest+bank%26quot%3b))+AND+((MH+%26quot%3bDeveloping+Countries%26quot%3b+OR+MH+%26quot%3blow+and+middle+income+countries%26quot%3b+OR+MH+%26quot%3bAfghanistan%26quot%3b+OR+MH+%26quot%3bAlbania%26quot%3b+OR+MH+%26quot%3bAlgeria%26quot%3b+OR+MH+%26quot%3bAngola%26quot%3b+OR+MH+%26quot%3bArgentina%26quot%3b+OR+MH+%26quot%3bArmenia%26quot%3b+OR+MH+%26quot%3bAzerbaijan%26quot%3b+OR+MH+%26quot%3bBangladesh%26quot%3b+OR+MH+%26quot%3bBelarus%26quot%3b+OR+MH+%26quot%3bBelize%26quot%3b+OR+MH+%26quot%3bBenin%26quot%3b+OR+MH+%26quot%3bBhutan%26quot%3b+OR+MH+%26quot%3bBolivia%26quot%3b+OR+MH+%26quot%3bBosnia-Herzegovina%26quot%3b+OR+MH+%26quot%3bBotswana%26quot%3b+OR+MH+%26quot%3bBrazil%26quot%3b+OR+MH+%26quot%3bBurkina+Faso%26quot%3b+OR+MH+%26quot%3bBurundi%26quot%3b+OR+MH+%26quot%3bCambodia%26quot%3b+OR+MH+%26quot%3bCameroon%26quot%3b+OR+MH+%26quot%3bCape+Verde%26quot%3b+OR+MH+%26quot%3bCentral+African+Republic%26quot%3b+OR+MH+%26quot%3bChad%26quot%3b+OR+MH+%26quot%3bChina%26quot%3b+OR+MH+%26quot%3bColombia%26quot%3b+OR+MH+%26quot%3bComoros%26quot%3b+OR+MH+%26quot%3bCongo%26quot%3b+OR+MH+%26quot%3bCosta+Rica%26quot%3b+OR+MH+%26quot%3bCote+d%26%2339%3bIvoire%26quot%3b+OR+MH+%26quot%3bCuba%26quot%3b+OR+MH+%26quot%3bDemocratic+Republic+of+Congo%26quot%3b+OR+MH+%26quot%3bDjibouti%26quot%3b+OR+MH+%26quot%3bDominica%26quot%3b+OR+MH+%26quot%3bDominican+Republic%26quot%3b+OR+MH+%26quot%3bEast+Timor%26quot%3b+OR+MH+%26quot%3bEcuador%26quot%3b+OR+MH+%26quot%3bEgypt%26quot%3b+OR+MH+%26quot%3bEl+Salvador%26quot%3b+OR+MH+%26quot%3bEquatorial+Guinea%26quot%3b+OR+MH+%26quot%3bEritrea%26quot%3b+OR+MH+%26quot%3bEthiopia%26quot%3b+OR+MH+%26quot%3bFiji%26quot%3b+OR+MH+%26quot%3bGabon%26quot%3b+OR+MH+%26quot%3bGambia%26quot%3b+OR+MH+%26quot%3bGeorgia+(Republic)%26quot%3b+OR+MH+%26quot%3bGhana%26quot%3b+OR+MH+%26quot%3bGuatemala%26quot%3b+OR+%26quot%3bMH+%26quot%3bGuinea%26quot%3b+or+MH+%26quot%3bGuinea-Bissau%26quot%3b+or+MH+%26quot%3bGuyana%26quot%3b+or+MH+%26quot%3bHaiti%26quot%3b+or+MH+%26quot%3bHonduras%26quot%3b+or+MH+%26quot%3bIndia%26quot%3b+or+MH+%26quot%3bIndian+Ocean+Islands%26quot%3b+or+MH+%26quot%3bIndonesia%26quot%3b+or+MH+%26quot%3bIran%26quot%3b+or+MH+%26quot%3bIraq%26quot%3b+or+MH+%26quot%3bJamaica%26quot%3b+or+MH+%26quot%3bJordan%26quot%3b+or+MH+%26quot%3bKazakhstan%26quot%3b+or+MH+%26quot%3bKenya%26quot%3b+or+MH+%26quot%3bKyrgyzstan%26quot%3b+or+MH+%26quot%3bLaos%26quot%3b+or+MH+%26quot%3bLebanon%26quot%3b+or+MH+%26quot%3bLesotho%26quot%3b+or+MH+%26quot%3bLiberia%26quot%3b+or+MH+%26quot%3bLibya%26quot%3b+or+MH+%26quot%3bMacedonia+(Republic)%26quot%3b+or+MH+%26quot%3bMadagascar%26quot%3b+or+MH+%26quot%3bMalawi%26quot%3b+or+MH+%26quot%3bMalaysia%26quot%3b+or+MH+%26quot%3bMali%26quot%3b+or+MH+%26quot%3bMauritania%26quot%3b+or+MH+%26quot%3bMexico%26quot%3b+or+MH+%26quot%3bMicronesia%26quot%3b+or+MH+%26quot%3bMoldova%26quot%3b+or+MH+%26quot%3bMongolia%26quot%3b+or+MH+%26quot%3bMorocco%26quot%3b+or+MH+%26quot%3bMozambique%26quot%3b+or+MH+%26quot%3bMyanmar%26quot%3b+or+MH+%26quot%3bNamibia%26quot%3b+or+MH+%26quot%3bNepal%26quot%3b+or+MH+%26quot%3bNicaragua%26quot%3b+or+MH+%26quot%3bNiger%26quot%3b+or+MH+%26quot%3bNigeria%26quot%3b+or+MH+%26quot%3bNorth+Korea%26quot%3b+or+MH+%26quot%3bPakistan%26quot%3b+or+MH+%26quot%3bPanama%26quot%3b+or+MH+%26quot%3bPapua+New+Guinea%26quot%3b+or+MH+%26quot%3bParaguay%26quot%3b+or+MH+%26quot%3bPeru%26quot%3b+or+MH+%26quot%3bPhilippines%26quot%3b+or+MH+%26quot%3bPolynesia%26quot%3b+or+MH+%26quot%3bRwanda%26quot%3b+or+MH+%26quot%3bSamoa%26quot%3b+or+MH+%26quot%3bSenegal%26quot%3b+or+MH+%26quot%3bSerbia%26quot%3b+or+MH+%26quot%3bSierra+Leone%26quot%3b+or+MH+%26quot%3bSomalia%26quot%3b+or+MH+%26quot%3bSouth+Africa%26quot%3b+or+MH+%26quot%3bSri+Lanka%26quot%3b+or+MH+%26quot%3bSudan%26quot%3b+or+MH+Suriname+or+MH+%26quot%3bSwaziland%26quot%3b+or+MH+%26quot%3bSyria%26quot%3b+or+MH+%26quot%3bTajikistan%26quot%3b+or+MH+%26quot%3bTanzania%26quot%3b+or+MH+%26quot%3bThailand%26quot%3b+or+MH+%26quot%3bTogo%26quot%3b+or+MH+%26quot%3bTunisia%26quot%3b+or+MH+%26quot%3bTurkey%26quot%3b+or+MH+%26quot%3bTurkmenistan%26quot%3b+or+MH+%26quot%3bUganda%26quot%3b+or+MH+%26quot%3bUkraine%26quot%3b+or+MH+%26quot%3bUzbekistan%26quot%3b+or+MH+%26quot%3bVenezuela%26quot%3b+or+MH+%26quot%3bVietnam%26quot%3b+or+MH+%26quot%3bYemen%26quot%3b+or+MH+%26quot%3bZambia%26quot%3b+or+MH+%26quot%3bZimbabwe)+OR+(TI+(%26quot%3bcabo+verde*%26quot%3b+OR+%26quot%3bdemocratic+people%26%2339%3bs+republic+of+korea%26quot%3b+OR+eswatini+OR+kiribati*+OR+%26quot%3bgaza+strip%26quot%3b+OR+grenada+OR+grenadian*+OR+grenadines+OR+kosov*+OR+%26quot%3blao+people%26%2339%3bs+democratic+republic%26quot%3b+OR+maldiv*+OR+%26quot%3bmarshall+island*%26quot%3b+OR+mauriti*+OR+montenegr*+OR+montserrat*+OR+nauru*+OR+niue*+OR+palestin*+OR+%26quot%3bsaint+helena*%26quot%3b+OR+%26quot%3bsaint+lucia*%26quot%3b+OR+%26quot%3bsaint+vincent*%26quot%3b+OR+%26quot%3bsao+tome*%26quot%3b+OR+%26quot%3bsolomon+island*%26quot%3b+OR+%26quot%3bsouth+sudan*%26quot%3b+OR+timorese+OR+tokelau*+OR+tonga*+OR+tuvalu*+OR+vanuatu*+OR+%26quot%3bwallis+and+futuna%26quot%3b+OR+%26quot%3bwest+bank%26quot%3b)+OR+AB+(%26quot%3bcabo+verde*%26quot%3b+OR+%26quot%3bdemocratic+people%26%2339%3bs+republic+of+korea%26quot%3b+OR+eswatini+OR+kiribati*+OR+%26quot%3bgaza+strip%26quot%3b+OR+grenada+OR+grenadian*+OR+grenadines+OR+kosov*+OR+%26quot%3blao+people%26%2339%3bs+democratic+republic%26quot%3b+OR+maldiv*+OR+%26quot%3bmarshall+island*%26quot%3b+OR+mauriti*+OR+montenegr*+OR+montserrat*+OR+nauru*+OR+niue*+OR+palestin*+OR+%26quot%3bsaint+helena*%26quot%3b+OR+%26quot%3bsaint+lucia*%26quot%3b+OR+%26quot%3bsaint+vincent*%26quot%3b+OR+%26quot%3bsao+tome*%26quot%3b+OR+%26quot%3bsolomon+island*%26quot%3b+OR+%26quot%3bsouth+sudan*%26quot%3b+OR+timorese+OR+tokelau*+OR+tonga*+OR+tuvalu*+OR+vanuatu*+OR+%26quot%3bwallis+and+futuna%26quot%3b+OR+%26quot%3bwest+bank%26quot%3b))))+OR+(MH+%26quot%3bBangladesh%26quot%3b+OR+MH+%26quot%3bBelarus%26quot%3b+OR+MH+%26quot%3bBelize%26quot%3b+OR+MH+%26quot%3bBenin%26quot%3b+OR+MH+%26quot%3bBhutan%26quot%3b+OR+MH+%26quot%3bBolivia%26quot%3b+OR+MH+%26quot%3bBosnia-Herzegovina%26quot%3b+OR+MH+%26quot%3bBotswana%26quot%3b+OR+MH+%26quot%3bBrazil%26quot%3b+OR+MH+%26quot%3bBurkina+Faso%26quot%3b+OR+MH+%26quot%3bBurundi%26quot%3b+OR+MH+%26quot%3bCambodia%26quot%3b+OR+MH+%26quot%3bCameroon%26quot%3b+OR+MH+%26quot%3bCape+Verde%26quot%3b+OR+MH+%26quot%3bCentral+African+Republic%26quot%3b+OR+MH+%26quot%3bChad%26quot%3b+OR+MH+%26quot%3bChina%26quot%3b+OR+MH+%26quot%3bColombia%26quot%3b+OR+MH+%26quot%3bComoros%26quot%3b+OR+MH+%26quot%3bCongo%26quot%3b+OR+MH+%26quot%3bCosta+Rica%26quot%3b+OR+MH+%26quot%3bCote+d%26%2339%3bIvoire%26quot%3b+OR+MH+%26quot%3bCuba%26quot%3b+OR+MH+%26quot%3bDemocratic+Republic+of+Congo%26quot%3b+OR+MH+%26quot%3bDjibouti%26quot%3b+OR+MH+%26quot%3bDominica%26quot%3b+OR+MH+%26quot%3bDominican+Republic%26quot%3b+OR+MH+%26quot%3bEast+Timor%26quot%3b+OR+MH+%26quot%3bEcuador%26quot%3b+OR+MH+%26quot%3bEgypt%26quot%3b+OR+MH+%26quot%3bEl+Salvador%26quot%3b+OR+MH+%26quot%3bEquatorial+Guinea%26quot%3b+OR+MH+%26quot%3bEritrea%26quot%3b+OR+MH+%26quot%3bEthiopia%26quot%3b+OR+MH+%26quot%3bFiji%26quot%3b+OR+MH+%26quot%3bGabon%26quot%3b+OR+MH+%26quot%3bGambia%26quot%3b+OR+MH+%26quot%3bGeorgia+(Republic)%26quot%3b+OR+MH+%26quot%3bGhana%26quot%3b+OR+MH+%26quot%3bGuatemala%26quot%3b+OR+%26quot%3bMH+%26quot%3bGuinea%26quot%3b+or+MH+%26quot%3bGuinea-Bissau%26quot%3b+or+MH+%26quot%3bGuyana%26quot%3b+or+MH+%26quot%3bHaiti%26quot%3b+or+MH+%26quot%3bHonduras%26quot%3b+or+MH+%26quot%3bIndia%26quot%3b+or+MH+%26quot%3bIndian+Ocean+Islands%26quot%3b+or+MH+%26quot%3bIndonesia%26quot%3b+or+MH+%26quot%3bIran%26quot%3b+or+MH+%26quot%3bIraq%26quot%3b+or+MH+%26quot%3bJamaica%26quot%3b+or+MH+%26quot%3bJordan%26quot%3b+or+MH+%26quot%3bKazakhstan%26quot%3b+or+MH+%26quot%3bKenya%26quot%3b+or+MH+%26quot%3bKyrgyzstan%26quot%3b+or+MH+%26quot%3bLaos%26quot%3b+or+MH+%26quot%3bLebanon%26quot%3b+or+MH+%26quot%3bLesotho%26quot%3b+or+MH+%26quot%3bLiberia%26quot%3b+or+MH+%26quot%3bLibya%26quot%3b+or+MH+%26quot%3bMacedonia+(Republic)%26quot%3b+or+MH+%26quot%3bMadagascar%26quot%3b+or+MH+%26quot%3bMalawi%26quot%3b+or+MH+%26quot%3bMalaysia%26quot%3b+or+MH+%26quot%3bMali%26quot%3b+or+MH+%26quot%3bMauritania%26quot%3b+or+MH+%26quot%3bMexico%26quot%3b+or+MH+%26quot%3bMicronesia%26quot%3b+or+MH+%26quot%3bMoldova%26quot%3b+or+MH+%26quot%3bMongolia%26quot%3b+or+MH+%26quot%3bMorocco%26quot%3b+or+MH+%26quot%3bMozambique%26quot%3b+or+MH+%26quot%3bMyanmar%26quot%3b+or+MH+%26quot%3bNamibia%26quot%3b+or+MH+%26quot%3bNepal%26quot%3b+or+MH+%26quot%3bNicaragua%26quot%3b+or+MH+%26quot%3bNiger%26quot%3b+or+MH+%26quot%3bNigeria%26quot%3b+or+MH+%26quot%3bNorth+Korea%26quot%3b+or+MH+%26quot%3bPakistan%26quot%3b+or+MH+%26quot%3bPanama%26quot%3b+or+MH+%26quot%3bPapua+New+Guinea%26quot%3b+or+MH+%26quot%3bParaguay%26quot%3b+or+MH+%26quot%3bPeru%26quot%3b+or+MH+%26quot%3bPhilippines%26quot%3b+or+MH+%26quot%3bPolynesia%26quot%3b+or+MH+%26quot%3bRwanda%26quot%3b+or+MH+%26quot%3bSamoa%26quot%3b+or+MH+%26quot%3bSenegal%26quot%3b+or+MH+%26quot%3bSerbia%26quot%3b+or+MH+%26quot%3bSierra+Leone%26quot%3b+or+MH+%26quot%3bSomalia%26quot%3b+or+MH+%26quot%3bSouth+Africa%26quot%3b+or+MH+%26quot%3bSri+Lanka%26quot%3b+or+MH+%26quot%3bSudan%26quot%3b+or+MH+Suriname+or+MH+%26quot%3bSwaziland%26quot%3b+or+MH+%26quot%3bSyria%26quot%3b+or+MH+%26quot%3bTajikistan%26quot%3b+or+MH+%26quot%3bTanzania%26quot%3b+or+MH+%26quot%3bThailand%26quot%3b+or+MH+%26quot%3bTogo%26quot%3b+or+MH+%26quot%3bTunisia%26quot%3b+or+MH+%26quot%3bTurkey%26quot%3b+or+MH+%26quot%3bTurkmenistan%26quot%3b+or+MH+%26quot%3bUganda%26quot%3b+or+MH+%26quot%3bUkraine%26quot%3b+or+MH+%26quot%3bUzbekistan%26quot%3b+or+MH+%26quot%3bVenezuela%26quot%3b+or+MH+%26quot%3bVietnam%26quot%3b+or+MH+%26quot%3bYemen%26quot%3b+or+MH+%26quot%3bZambia%26quot%3b+or+MH+%26quot%3bZimbabwe))+AND+((%26quot%3bsexual+health%26quot%3b+OR+HIV+OR+%26quot%3bsexually+transmitted+disease%26quot%3b+OR+%26quot%3bsexual+behaviour%26quot%3b+OR+%26quot%3bfamily+planning%26quot%3b+OR+%26quot%3breproductive+health%26quot%3b+OR+abortion+OR+%26quot%3bsexual+violence%26quot%3b+OR+contraception+OR+%26quot%3bsexual+identity%26quot%3b+OR+%26quot%3bsexual+dysfunction%26quot%3b+OR+%26quot%3bpregnancy%26quot%3b+OR+%26quot%3bmotherhood%26quot%3b)+AND+(%26quot%3bSevere+mental+illness%26quot%3b+OR+schizophrenia+OR+bipolar+OR+psychosis)+AND+(((TI+(%26quot%3bcabo+verde*%26quot%3b+OR+%26quot%3bdemocratic+people%26%2339%3bs+republic+of+korea%26quot%3b+OR+eswatini+OR+kiribati*+OR+%26quot%3bgaza+strip%26quot%3b+OR+grenada+OR+grenadian*+OR+grenadines+OR+kosov*+OR+%26quot%3blao+people%26%2339%3bs+democratic+republic%26quot%3b+OR+maldiv*+OR+%26quot%3bmarshall+island*%26quot%3b+OR+mauriti*+OR+montenegr*+OR+montserrat*+OR+nauru*+OR+niue*+OR+palestin*+OR+%26quot%3bsaint+helena*%26quot%3b+OR+%26quot%3bsaint+lucia*%26quot%3b+OR+%26quot%3bsaint+vincent*%26quot%3b+OR+%26quot%3bsao+tome*%26quot%3b+OR+%26quot%3bsolomon+island*%26quot%3b+OR+%26quot%3bsouth+sudan*%26quot%3b+OR+timorese+OR+tokelau*+OR+tonga*+OR+tuvalu*+OR+vanuatu*+OR+%26quot%3bwallis+and+futuna%26quot%3b+OR+%26quot%3bwest+bank%26quot%3b)+OR+AB+(%26quot%3bcabo+verde*%26quot%3b+OR+%26quot%3bdemocratic+people%26%2339%3bs+republic+of+korea%26quot%3b+OR+eswatini+OR+kiribati*+OR+%26quot%3bgaza+strip%26quot%3b+OR+grenada+OR+grenadian*+OR+grenadines+OR+kosov*+OR+%26quot%3blao+people%26%2339%3bs+democratic+republic%26quot%3b+OR+maldiv*+OR+%26quot%3bmarshall+island*%26quot%3b+OR+mauriti*+OR+montenegr*+OR+montserrat*+OR+nauru*+OR+niue*+OR+palestin*+OR+%26quot%3bsaint+helena*%26quot%3b+OR+%26quot%3bsaint+lucia*%26quot%3b+OR+%26quot%3bsaint+vincent*%26quot%3b+OR+%26quot%3bsao+tome*%26quot%3b+OR+%26quot%3bsolomon+island*%26quot%3b+OR+%26quot%3bsouth+sudan*%26quot%3b+OR+timorese+OR+tokelau*+OR+tonga*+OR+tuvalu*+OR+vanuatu*+OR+%26quot%3bwallis+and+futuna%26quot%3b+OR+%26quot%3bwest+bank%26quot%3b))+AND+((MH+%26quot%3bDeveloping+Countries%26quot%3b+OR+MH+%26quot%3blow+and+middle+income+countries%26quot%3b+OR+MH+%26quot%3bAfghanistan%26quot%3b+OR+MH+%26quot%3bAlbania%26quot%3b+OR+MH+%26quot%3bAlgeria%26quot%3b+OR+MH+%26quot%3bAngola%26quot%3b+OR+MH+%26quot%3bArgentina%26quot%3b+OR+MH+%26quot%3bArmenia%26quot%3b+OR+MH+%26quot%3bAzerbaijan%26quot%3b+OR+MH+%26quot%3bBangladesh%26quot%3b+OR+MH+%26quot%3bBelarus%26quot%3b+OR+MH+%26quot%3bBelize%26quot%3b+OR+MH+%26quot%3bBenin%26quot%3b+OR+MH+%26quot%3bBhutan%26quot%3b+OR+MH+%26quot%3bBolivia%26quot%3b+OR+MH+%26quot%3bBosnia-Herzegovina%26quot%3b+OR+MH+%26quot%3bBotswana%26quot%3b+OR+MH+%26quot%3bBrazil%26quot%3b+OR+MH+%26quot%3bBurkina+Faso%26quot%3b+OR+MH+%26quot%3bBurundi%26quot%3b+OR+MH+%26quot%3bCambodia%26quot%3b+OR+MH+%26quot%3bCameroon%26quot%3b+OR+MH+%26quot%3bCape+Verde%26quot%3b+OR+MH+%26quot%3bCentral+African+Republic%26quot%3b+OR+MH+%26quot%3bChad%26quot%3b+OR+MH+%26quot%3bChina%26quot%3b+OR+MH+%26quot%3bColombia%26quot%3b+OR+MH+%26quot%3bComoros%26quot%3b+OR+MH+%26quot%3bCongo%26quot%3b+OR+MH+%26quot%3bCosta+Rica%26quot%3b+OR+MH+%26quot%3bCote+d%26%2339%3bIvoire%26quot%3b+OR+MH+%26quot%3bCuba%26quot%3b+OR+MH+%26quot%3bDemocratic+Republic+of+Congo%26quot%3b+OR+MH+%26quot%3bDjibouti%26quot%3b+OR+MH+%26quot%3bDominica%26quot%3b+OR+MH+%26quot%3bDominican+Republic%26quot%3b+OR+MH+%26quot%3bEast+Timor%26quot%3b+OR+MH+%26quot%3bEcuador%26quot%3b+OR+MH+%26quot%3bEgypt%26quot%3b+OR+MH+%26quot%3bEl+Salvador%26quot%3b+OR+MH+%26quot%3bEquatorial+Guinea%26quot%3b+OR+MH+%26quot%3bEritrea%26quot%3b+OR+MH+%26quot%3bEthiopia%26quot%3b+OR+MH+%26quot%3bFiji%26quot%3b+OR+MH+%26quot%3bGabon%26quot%3b+OR+MH+%26quot%3bGambia%26quot%3b+OR+MH+%26quot%3bGeorgia+(Republic)%26quot%3b+OR+MH+%26quot%3bGhana%26quot%3b+OR+MH+%26quot%3bGuatemala%26quot%3b+OR+%26quot%3bMH+%26quot%3bGuinea%26quot%3b+or+MH+%26quot%3bGuinea-Bissau%26quot%3b+or+MH+%26quot%3bGuyana%26quot%3b+or+MH+%26quot%3bHaiti%26quot%3b+or+MH+%26quot%3bHonduras%26quot%3b+or+MH+%26quot%3bIndia%26quot%3b+or+MH+%26quot%3bIndian+Ocean+Islands%26quot%3b+or+MH+%26quot%3bIndonesia%26quot%3b+or+MH+%26quot%3bIran%26quot%3b+or+MH+%26quot%3bIraq%26quot%3b+or+MH+%26quot%3bJamaica%26quot%3b+or+MH+%26quot%3bJordan%26quot%3b+or+MH+%26quot%3bKazakhstan%26quot%3b+or+MH+%26quot%3bKenya%26quot%3b+or+MH+%26quot%3bKyrgyzstan%26quot%3b+or+MH+%26quot%3bLaos%26quot%3b+or+MH+%26quot%3bLebanon%26quot%3b+or+MH+%26quot%3bLesotho%26quot%3b+or+MH+%26quot%3bLiberia%26quot%3b+or+MH+%26quot%3bLibya%26quot%3b+or+MH+%26quot%3bMacedonia+(Republic)%26quot%3b+or+MH+%26quot%3bMadagascar%26quot%3b+or+MH+%26quot%3bMalawi%26quot%3b+or+MH+%26quot%3bMalaysia%26quot%3b+or+MH+%26quot%3bMali%26quot%3b+or+MH+%26quot%3bMauritania%26quot%3b+or+MH+%26quot%3bMexico%26quot%3b+or+MH+%26quot%3bMicronesia%26quot%3b+or+MH+%26quot%3bMoldova%26quot%3b+or+MH+%26quot%3bMongolia%26quot%3b+or+MH+%26quot%3bMorocco%26quot%3b+or+MH+%26quot%3bMozambique%26quot%3b+or+MH+%26quot%3bMyanmar%26quot%3b+or+MH+%26quot%3bNamibia%26quot%3b+or+MH+%26quot%3bNepal%26quot%3b+or+MH+%26quot%3bNicaragua%26quot%3b+or+MH+%26quot%3bNiger%26quot%3b+or+MH+%26quot%3bNigeria%26quot%3b+or+MH+%26quot%3bNorth+Korea%26quot%3b+or+MH+%26quot%3bPakistan%26quot%3b+or+MH+%26quot%3bPanama%26quot%3b+or+MH+%26quot%3bPapua+New+Guinea%26quot%3b+or+MH+%26quot%3bParaguay%26quot%3b+or+MH+%26quot%3bPeru%26quot%3b+or+MH+%26quot%3bPhilippines%26quot%3b+or+MH+%26quot%3bPolynesia%26quot%3b+or+MH+%26quot%3bRwanda%26quot%3b+or+MH+%26quot%3bSamoa%26quot%3b+or+MH+%26quot%3bSenegal%26quot%3b+or+MH+%26quot%3bSerbia%26quot%3b+or+MH+%26quot%3bSierra+Leone%26quot%3b+or+MH+%26quot%3bSomalia%26quot%3b+or+MH+%26quot%3bSouth+Africa%26quot%3b+or+MH+%26quot%3bSri+Lanka%26quot%3b+or+MH+%26quot%3bSudan%26quot%3b+or+MH+Suriname+or+MH+%26quot%3bSwaziland%26quot%3b+or+MH+%26quot%3bSyria%26quot%3b+or+MH+%26quot%3bTajikistan%26quot%3b+or+MH+%26quot%3bTanzania%26quot%3b+or+MH+%26quot%3bThailand%26quot%3b+or+MH+%26quot%3bTogo%26quot%3b+or+MH+%26quot%3bTunisia%26quot%3b+or+MH+%26quot%3bTurkey%26quot%3b+or+MH+%26quot%3bTurkmenistan%26quot%3b+or+MH+%26quot%3bUganda%26quot%3b+or+MH+%26quot%3bUkraine%26quot%3b+or+MH+%26quot%3bUzbekistan%26quot%3b+or+MH+%26quot%3bVenezuela%26quot%3b+or+MH+%26quot%3bVietnam%26quot%3b+or+MH+%26quot%3bYemen%26quot%3b+or+MH+%26quot%3bZambia%26quot%3b+or+MH+%26quot%3bZimbabwe)+OR+(TI+(%26quot%3bcabo+verde*%26quot%3b+OR+%26quot%3bdemocratic+people%26%2339%3bs+republic+of+korea%26quot%3b+OR+eswatini+OR+kiribati*+OR+%26quot%3bgaza+strip%26quot%3b+OR+grenada+OR+grenadian*+OR+grenadines+OR+kosov*+OR+%26quot%3blao+people%26%2339%3bs+democratic+republic%26quot%3b+OR+maldiv*+OR+%26quot%3bmarshall+island*%26quot%3b+OR+mauriti*+OR+montenegr*+OR+montserrat*+OR+nauru*+OR+niue*+OR+palestin*+OR+%26quot%3bsaint+helena*%26quot%3b+OR+%26quot%3bsaint+lucia*%26quot%3b+OR+%26quot%3bsaint+vincent*%26quot%3b+OR+%26quot%3bsao+tome&type=1&searchMode=Standard&site=ehost-live

PsycInfo

<https://ovidsp.ovid.com/ovidweb.cgi?T=JS&NEWS=N&PAGE=main&SHAREDSEARCHID=5HDUVsN63MI3lT8WU1f3gXIBR8rT8C3chrGZzsL9DxR8j52hhCSLZOxQKXpRgqMoJ>

1

exp Sexual Attitudes/ or exp Psychosexual Behavior/ or exp Sexual Satisfaction/ or exp Sexual Function Disturbances/ or exp Sexual Health/ or exp Marital Relations/ or exp Sexuality/ or sexual health.mp.

00:01

2

family planning.mp. or exp Family Planning/

00:01

3

exp HIV/ or exp Sexually Transmitted Diseases/ or exp Sexual Risk Taking/ or exp Condoms/ or sexually transmitted infection.mp. or exp Sex Work/

00:01

4

reproductive health.mp. or exp Reproductive Health/

00:01

5

exp Sexual Abuse/ or exp Rape/ or exp Sex Offenses/ or sexual violence.mp. or exp Intimate Partner Violence/

00:01

6

contraception.mp. or exp Birth Control/

00:01

7

abortion.mp.

8

exp Psychosexual Behavior/ or sexual identity.mp. or exp Sexuality/

9

exp Female Sexual Dysfunction/ or exp Psychosexual Behavior/ or sexual dysfunction.mp.

10

exp Pregnancy/ or pregnancy.mp.

11

motherhood.mp.

12

severe mental illness.mp. or exp Serious Mental Illness/

13

schizophrenia.mp. or exp Schizophrenia/

14

bipolar disorder.mp. or exp Bipolar Disorder/

15

exp Psychosis/ or psychosis.mp. or exp Postpartum Psychosis/

16

developing countries.mp. or exp Developing Countries/

17

(low middle income countr* or "low* and middle income countr*" or "low* or middle income countr*" or lmic*).ti,ab.

18

(afghan* or albania* or algeria* or angola* or argentin* or armenia* or azerbaijan* or azeri* or bangladesh* or basotho* or belarus* or belize* or benin* or bhutan* or bissau-guinea* or bolivia* or bosnia* or botswan* or brazil* or burkina faso or burkinabe* or burmese or burundi* or cabo verde* or cambodia* or cameroon* or central african republic or chad or chadian* or china or chinese or columbia* or comor* or congo* or costa rica* or cote d'ivoire or cuba* or democratic people's republic of korea or djibouti* or dominica* or ecuador* or egypt* or el salvador* or equatoguinea* or equatorial guinea* or eritrea* or eswatini or ethiopia* or fiji* or filipin* or futunan* or gabon* or gambia* or gaza strip or georgia* or ghana* or grenada or grenadian* or grenadines or guatemala* or guinea-bissau* or guyan* or haiti* or herzegovin* or hondura* or india or indian* or indonesia* or iran* or iraq* or ivorian* or ivory coast or i-kiribati* or jamaica* or jordan* or kazakh* or kenya* or kirghiz* or kirgiz* or kiribati* or kosov* or kyrgyz* or lao people's democratic republic or lao* or leban* or lesotho* or liberia* or libyan* or madagasca* or malawi* or malaysia* or maldiv* or mali or malian* or malinese* or marshall island* or marshallese* or mauritania* or mauriti* or mexican* or mexico or micronesia* or moldova* or mongolia* or mongol or mongol's or mongols* or montenegr* or montserrat* or morocc* or mosotho* or mozambique or mozambican* or myanma* or nepal* or namibia* or nauru* or nicaragua* or niue* or niger* or ni-vanuatu* or north korea* or macedonia* or pakistan* or palestin* or panama* or papua new guinea* or papuan* or paraguay* or peru* or philippine* or rwanda* or saint helen* or saint lucia* or saint vincent* or salvadoran* or salvadorean* or salvadorian* or samoa* or sao tome* or senegal* or serbia* or sierra leone* or solomon island* or somali* or south africa* or sri lanka* or sudan* or suriname* or swati* or swazi* or syria* or tajik* or tanzania* or thai* or timor-leste or timorese or togo or togolese or tokelau* or tonga* or tunisia* or turk or turkey or turkish or turkmen* or turk's or turks or tuvalu* or uganda* or ukrain* or uzbek* or vanuatu* or venezuela* or vietnam* or "wallis and fortuna" or wallisian* or west bank or yemen* or zambia* or zimbabwe*).ti,ab.

19

1 or 2 or 3 or 4 or 5 or 6 or 7 or 8 or 9 or 10 or 11

20

12 or 13 or 14 or 15

21

16 or 17 or 18

22

19 and 20 and 21

results 898- duplicates removed 893

Screening

MEDLINE + EMBASE = 347 duplicates

MEDLINE + EMBASE + psycinfo = 562 duplicates

MEDLINE + EMBASE + psycinfo + CINAHL = 603 duplicates

After duplicates removed 2730 studies

After title and abstract screening 280 studies

After full text screening

07/07/23 MEDLINE search rerun and results for 2022-2024 included only. 78 studies identified. 8 studies fit inclusion criteria. after duplicates removed, 2 studies:

10.4102/sajpsychiatry.v29i0.1918 Mwelase 2023 **HIV** prevalence **and** access to **HIV** testing **and** care in patients with psychosis in **South** **Africa**. (I)

10.1016/j.pecinn.2022.100016 Yahyavi 2022 Sex education for patients with **severe** **mental** **illness** in **Iran**: A qualitative study (E- not specific to women)

One study added.
