## Appendix 2 Supplementary tables for "Sexual and Reproductive Health Needs of Women with Severe Mental Illness in Low- and Middle-Income Countries: A Scoping Review"

### Table 1.1.1. HIV Prevalence: Quantitative Studies

| Author, year | Country | HIV prevalence | |
| --- | --- | --- | --- |
|  |  | **Women with SMI** | **Comparison** |
| Aboobaker, 2022 [1] | South Africa | 55/181 (30%) | 14/187 (7%) Men with SMI |
| Carey, 2007[2] | India | 7/375 (1.9%) | 9/573 (1.6%) Men with SMI |
| Collins, 2009 [3] | South Africa | 33.3% | 19.7% Men with SMI |
| Guimarães, 2014[4] | Brazil | 8/1161 (0.69%) | 7/1076 (0.65%) Men with SMI |
| Henning, 2012[5] | South Africa | 16/95 (17%) | 8/100 (8%) Men with SMI |
| Lundberg, 2013[6] | Uganda | 49/343 (14.3%) | 19/259 (7.3%) Men with SMI |
| Madziro‑Ruwizhu, 2019[7] | Zimbabwe | 29/128 (22.7%) | 10/142 (7%) Men with SMI |
| Maling, 2011[8] | Uganda | 35/151 (23%) | 15/171(9%) Men with SMI |
| Mashaphu, 2007 [9] | South Africa | 6/13 (46.2%) | 9/50 (18%) Men with SMI |
| Mere, 2018[10] | South Africa | 4/121 (61%) | 26/80 (33%) men with SMI  *note this was not a random sample |
| Mpango, 2022 [11] | Uganda | OR 2.67 (CI 1.61; 4.43) | OR 1 Men with SMI |
| Mwelase, 2023 [12] | South Africa | 37/94 (39%) | 25/202 (12%) Men with SMI |
| Opondo, 2008[13] | Botswana | 187/356 (53%) | 112/599 (19%) Men with SMI |
| Singh, 2009[14] | South Africa | 36/88 (40.1%) | 20.7% Black Women aged 15-45 in the general population in South Africa |

### Table 1.2.1 HIV Risk Behaviour: Quantitative Studies

| Author, year | Country | HIV Risk Behaviour | |
| --- | --- | --- | --- |
|  |  | **Women with SMI** | **Comparison** |
| Abayomi, 2013[15] | Nigeria | 13/28 (46.4%) had high risk sexual behaviour^[1]^ | 36/74 (48.6%) men had high risk sexual behaviour¹ |
| Chandra, 2003 (a) [16] | India | 14% had lifetime high risk sexual behaviour in the HIV risk screening instrument | No comparison |
| Chandra, 2003 (c) [17] | India | 46% reported multiple sexual partners (not significant)  27% reported exchanging sex for money (p=<0.05)  0% reported exchanging money for sex (p=<0.001) | 65% of men with SMI  9% men with SMI  66% men with SMI |
| Chopra, 1998[18] | India | 14/30 (47%) women had high risk heterosexual intercourse¹ | 13/29 (49%) men had high risk heterosexual intercourse¹ |
| Gebeyehu, 2021[19] | Ethiopia | 48/113 (42%) women risky sexual behaviour¹ | 63/110 men (57%) men had risky sexual behaviour¹ |
| Guimarães, 2010[20] | Brazil | 83.6% women had unprotected sex at least once in their lifetime¹ | 76.9% men had unprotected sex at least once in their lifetime¹ |
| Lundberg, 2015[21] | Uganda | 13/343 (3.8%) of women had multiple sexual partners¹  11/343 (3.2%) of women had multiple partners with no condom use ¹ | 54/257 (21%) of men had multiple sexual partners¹  54/257 (21%) of men had multiple partners with no condom use ¹ |
| Marengo, 2015 (b)[22] | Argentina | 21.4% of women with bipolar disorder (WBD) had casual partners; 14.5% WBD had multiple partners; 37.5% of WBD had non-monogamous sexual partners; 14.3% of WBD had partners with unknown HIV status; 42.9% WBD had consistent condom use; 33% WBD used alcohol or drugs before sex. | The control group were women without bipolar disorder. 5.4% of controls had casual partners; 8.3% controls had multiple partners; 17.9% of controls had non-monogamous sexual partners; 3.6% controls had partners with unknown HIV status; 40.4% controls had consistent condom use; 20% controls used alcohol or drugs before sex. |
| Negash, 2019[23] | Uganda | 22/155 (14%) women had risky sexual behaviour on health survey for assessment of risky sexual behaviour among HIV/AIDS patients (adapted for local context) | 147/274 (54%) men had risky sexual behaviour on health survey for assessment of risky sexual behaviour among HIV/AIDS patients (adapted for local context) |
| Obo, 2019 [24] | Ethiopia | 40/124 (32%) women had risky sexual behaviour in the Behavioural surveillance survey (adapted) | 107/173 (62%) men had risky sexual behaviour in the Behavioural surveillance survey (adapted) |
| Peixoto, 2014[25] | Brazil | 88.8% women reported unprotected sex in the last six months ¹ | 77.2% men reported unprotected sex in the last six months ¹ |
| Tomruk 2006 [26] | Turkey | Women with schizophrenia: 10% had more than 1 partner, 6% multiple partners simultaneously, 4% IV substance addicted partners, 10% had intercourse with someone they knew less than a day, 10% had intercourse under influence of alcohol/drugs. Women with BD: 8% had multiple sexual partners, 4% had casual sex with stranger, 4% had anal intercourse | No comparison |
| Wainberg, 2008[27] | Brazil | 60.5% of the 16 sexually active women who reported not using a condom said this was due to their partners' preference. 31.3% said condoms weren’t available at the time of intercourse. Assessed using the Sexual Risk Behaviour Assessment Schedule | 50% of the 16 sexually active men who reported not using condoms cited ‘trust in their partner’, 18.8% said this was as they perceived themselves not to be at risk of HIV and 18.8% said it was due to partner preference. |

### Table 1.2.2. HIV Risk Behaviour: Qualitative Studies

| Author, year | Country | HIV Risk Behaviour | |
| --- | --- | --- | --- |
|  |  | **Study Aim & Population** | **Qualitative Findings: Women with SMI** |
| Collins, 2001[28] | South Africa | Study aim: This paper presents data from a qualitative study of South African mental health care providers, examining their perceptions of the HIV risk and experiences of sexuality among women with SMI.  Population: Men and women who had direct contact and clinical or rehabilitation responsibilities for individuals with mental illness or administrative duties in provincial mental health care departments. | More concern about pregnancy avoidance than HIV avoidance in an ethnographic study |
| Pinto, 2007 [29] | Brazil | Study aim: to Investigate the beliefs, values, attitudes, and motivations related to behaviours that would provide basis for developing interventions for prevention of HIV among male and female mental health service users  Population: People with SMI and admin and patient facing staff | Staff in a mental healthcare facility noted instances of unprotected sexual intercourse among patients and perceived them to be at a higher risk of HIV/STIs |
| Shahvari, 2020[30] | Iran | Study aim: to explore factors which should be considered in designing a sexuality education package for severe mental illness.  Population: SMI patients, their family, clinical staff in a psychiatric unit and a general physician | Some service providers believed that severe psychiatric patients are not able to resist the temptation to engage in risky behaviour. |

### Table 1.3.1. HIV Knowledge: Quantitative Studies

| Author, year | Country | HIV Knowledge | |
| --- | --- | --- | --- |
|  |  | **Women with SMI** | **Comparison** |
| Chandra, 2006[31] | India | Mean HIV knowledge score pre HIV- risk reduction education intervention 4.37, post intervention 9.47, 5 days post intervention 9.05 | Mean HIV knowledge score (men with SMI) pre HIV- risk reduction education intervention 7.85 men; post intervention 12.55; 5 days post intervention 11.85 |
| Chopra, 1998[18] | India | 24/30 (80%) of women had inadequate knowledge about HIV high risk behaviour | 15/29 (52%) of men with SMI had inadequate knowledge about HIV high risk behaviour |
| Matshoba, 2021[32] | South Africa | There was no significant difference in the HIV knowledge, HIV attitudes or HIV practices score between men and women with SMI | There was no significant difference in the HIV knowledge, HIV attitudes or HIV practices score between men and women with SMI |
| Melo, 2010[33] | Brazil | Women had a significantly higher HIV knowledge score than men (actual figures not reported) | Men had a significantly lower HIV knowledge score than women (actual figures not reported) |
| Peixoto, 2014[25] | Brazil | No significant difference between men and women in HIV/AIDS knowledge | No comparison |
| Tomruk, 2006[26] | Turkey | 46% WBD and 64% schizophrenia agreed that condoms are protective against AIDS;  58% WBD and 46% schizophrenia agreed that AIDS is frequent in IVDU | The control group were women without bipolar disorder. 42% control agreed that condoms are protective against AIDS; 76% control agreed that AIDS is frequent in IVDU |

### Table 1.4.1 Other Relevant Findings: Quantitative Studies

| Author, year | Country | Other Relevant Findings | |
| --- | --- | --- | --- |
|  |  | **Women with SMI** | **Comparison** |
| Joska, 2014[34] | South Africa | 34/85 (40%) women attended their HIV appointment within 6 months | 3/15 (20%) men attended their HIV appointment within 6 months |
| Souto, 2011[35] | Brazil | 350/1232 had HIV testing ever. No significant difference between women and men. | 350/1232 had HIV testing ever. No significant difference between women and men. |
| Data collected using a bespoke questionnaire rather than a validated tool | | | |

### Table 2.1. Sexual Function: Quantitative Studies

| Author, year | Country | Sexual Function | |  |
| --- | --- | --- | --- | --- |
|  |  | **Women with SMI** | **Comparison** | **Scale Used** |
| Ben, 2013[36] | Tunisia | Female gender was significantly correlated with sexual dysfunction | Male gender was not significantly correlated with sexual dysfunction | Sexual Behaviour Questionnaire (SBQ) |
| Bram, 2014[37] | Tunisia | 7 (43.7%) women experienced sexual dysfunction | 9 (40.9%) men experienced sexual dysfunction | Changes in Sexual Functioning Questionnaire (CSFQ) |
| Caqueo-Urizar, 2018[38] | Bolivia, Peru and Chile | Women had a higher sentimental life score (measuring satisfaction with love life) of 48.3 | Men had a lower sentimental life score (measuring satisfaction with love life) of 43.6 | Schizophrenia Quality of Life Questionnaire (SQoL18) |
| Dogu, 2012[39] | Turkey | 33/63 (52.4%) of women had a problem in their sexual life.  29/63 (46%) of women considered that their sexual problems are due to their illness.  53/63 (84.1%) of women have had no information about the influence of the illness and medications on their sexual functions.  56/63 (88.9%) of women said that their psychiatrists' did not ask them about their sexual life or about their sexual problems, and also didn't inform them about those topics. | 22/57 (38.6%) of men had a problem in their sexual life.  28/57 (49.1%) of men considered that their sexual problems are due to their illness.  45/57 (78.9%) of men have had no information about the influence of the illness and medications on their sexual functions.  44/57 (77.2%) of men said that their psychiatrists' did not ask them about their sexual life or about their sexual problems, and also didn't inform them about those topics. | Bespoke |
| Esan, 2018[40] | Nigeria | 19/45 (42.2%) women experienced some level of sexual dysfunction | 14/45 (31.1%) men experienced some level of sexual dysfunction | Arizona Sexual Experiences Questionnaire (ASEX) |
| Fanta, 2018[41] | Ethiopia | 78.6% prevalence of sexual dysfunction in women (number not reported) | 84.5% prevalence of sexual dysfunction in men (number not reported) | CSFQ |
| Halouani, 2018[42] | Tunisia | 81.3% prevalence of sexual dysfunction | Women without SMI had significantly less sexual dysfunction | Female Sexual Functioning Index (FSFI) |
| Hariri, 2009[43] | Turkey | Women with schizophrenia had a total GRISS score of 5.1 (a GRISS score over 5 shows sexual dysfunction)  Women with bipolar disorder had a total GRISS score of 4.6 (a GRISS score over 5 shows sexual dysfunction) | Men with schizophrenia had a total GRISS score of 5.1 (a GRISS score over 5 shows sexual dysfunction)  Men with bipolar disorder had a total GRISS score of 5.7 (a GRISS score over 5 shows sexual dysfunction) | Golombok Rust Inventory of Sexual Satisfaction (22.9) |
| Hocaoglu, 2014[44] | Turkey | 68% prevalence of sexual dysfunction in women | 46% prevalence of sexual dysfunction in men | ASEX |
| Hou, 2016[45] | China | 80.6% prevalence of sexual dysfunction in women | 60.7% prevalence of sexual dysfunction in men | ASEX |
| Huang, 2019[46] | China | 82.1% prevalence of sexual dysfunction in sexually active women. 82.7% in non sexually active women. | 67.8% prevalence of sexual dysfunction in sexually active men. 64.5% in non sexually active men. | ASEX |
| Incedere, 2017[47] | Turkey | Average ASEX score was 20.14 for women | Average ASEX score was 14.82 for women | ASEX |
| Kazour, 2020[48] | Lebanon | SBQ score for women who masturbate was 19.41  SBQ score for women who do not masturbate was 4.39 | SBQ score 21.37 for men who masturbate  SBQ score 3.24 for men who do not masturbate | SBQ |
| Kesebir, 2014 [49] | Turkey | ASEX female: 9.2 lithium 18.5 quetiapine 15.3 olanzapine  GRISS female: 43.8 lithium 32.2 quetiapine 35.7 olanzapine | ASEX male: 11.2 lithium 15.3 quetiapine 11.6 olanzapine  GRISS male: 72.6 lithium 87.3 quetiapine 84.2 olanzapine | ASEX and GRISS |
| Kumar, 2021 [50] | India | Sexual dysfunction was present in 24/29 (82.8%) of women taking olanzapine  Sexual dysfunction was present in 26/28 (92.9%) of women taking risperidone | No comparison | CSFQ |
| Nakhli, 2014[51] | Tunisia | Women had a higher ASEX total score (exact numbers not provided) | Men had a lower ASEX total score (exact numbers not provided) | ASEX |
| Olisah 2016[52] | Nigeria | 58.8% of women had sexual dysfunction | 40.2% of men had sexual dysfunction | International Index of Erectile Function (IIEF) Questionnaire for the male participants and the FSFI for the female participants. |
| Simiyon, 2006[53] | India | 70% of women had sexual dysfunction | No comparison | FSFI |
| Souaiby, 2020 [54] | Lebanon | 8/13 (61.5%) women had sexual dysfunction | 47/82 (57.3%) of men had sexual dysfunction | Psychotropic-Related Sexual Dysfunction Questionnaire (PRSexDQ) |
| Tharoor, 2015[55] | India | 19/67 (28.4%) women reported sexual dysfunction after taking antipsychotics | 29/69 (42%) men reported sexual dysfunction after taking antipsychotics | Psychotropic Related Sexual Dysfunction Questionnaire (PRSexDQ‐Salsex) |
| Wainberg, 2008[27] | Brazil | 28.6% of women experienced lack of interest in sexual activity. | 16.7% of men experienced lack of interest in sexual activity. | Sexual Risk Behavior Assessment Schedule (SERBAS) |
| Zhang, 2018 (b) [56] | China | 39/63 (62%) women had sexual dysfunction | 22/55 (40%) men had sexual dysfunction | ASEX |

### Table 2.2. Sexual Function: Qualitative Studies

| Author, year | Country | Sexual Function | |
| --- | --- | --- | --- |
|  |  | **Study Aim & Population** | **Qualitative Findings: Women with SMI** |
| Collins, 2001[28] | South Africa | Study aim: This paper presents data from a qualitative study of South African mental health care providers, examining their perceptions of the HIV risk and experiences of sexuality among women with SMI.  Population: Men and women who had direct contact and clinical or rehabilitation responsibilities for individuals with mental illness or administrative duties in provincial mental health care departments. | Almost all providers believed mental illness would affect women’s sexual life (the amount of sexual activity, kinds of sexual acts, and qualitative experience of sexuality). The clinicians’ perceptions of whether or not a patient was sexually active appeared to be related to the severity of psychiatric symptoms. Some viewed sexual activity in the women with psychiatric illness as qualitatively different from “normal” sexuality. Some providers viewed people with mental illness as having the same sexual desires as people without mental illness. Some providers acknowledged the stigmatizing impact of clinicians’ attitudes toward sexuality among women with mental illness. |
| Lundberg, 2012[57] | Uganda | Study aim: To understand how having a SMI may influence sexual risk behaviours and sexual health risks in a low-income sub-Saharan African country with high HIV prevalence.  Population: people with SMI | A female participant with bipolar disorder and HIV explained she had sex with several casual partners because ‘she wanted to die’, suggesting that casual sex was a self-destructive behaviour for this woman. A female participant with bipolar disorder suggested that during illness episodes she may be more likely to make impulsive decisions about sex. |
| Mirsepassi, 2022[58] | Iran | Study aim: to explore family knowledge about sexual health in patients with severe mental illness in Iran.  Population: people with SMI, their families, psychiatric staff and a general practitioner. | Common myths around sexual health in patients with SMI (codes: The lack of sexual needs in patients’, ‘feasibility of lifelong abstinence’, ‘restriction the only way of sexual health’) |
| Pinto, 2007 [29] | Brazil | Study aim: to Investigate the beliefs, values, attitudes, and motivations related to behaviours that would provide basis for developing interventions for prevention of HIV among male and female mental health service users  Population: People with SMI and admin and patient facing staff | Sexuality and all things related considered 'taboo' and 'forbidden' by 3 women with SMI. Professionals viewed sexual activity in people with SMI as being 'void of affection', described as 'animalistic' and of patients being highly 'oversexed'. Differing professional views: some saw sexual desire as being a consequence of the psychiatric condition and others an integral aspect of their lives and wellbeing. Some professionals viewed women as being more sexually aggressive compared to men, requiring professionals to intervene. Some described a myth that men with SMI are more sexually driven than female patients but did not find this myth to be true in their experience of working with patients with SMI. |
| Rezaie, 2020[59] | Iran | Study aim: to determine the unique post-discharge needs of Iranian women diagnosed with severe mental illness.  Population: people with SMI | Study participants expressed a particular need for information about women's health issues. They identified pregnancy, sex drive and effects of medication on the developing foetus as key topics of concern. Broadly, participants requested more information on the relationship between SMI and feminine identity. Study participants also stated that they want to know about the impact of SMI and medication on their sexual relationships. They expressed a desire to have a normal sexual relationship with their husbands. |
| Tumwakire, 2022[60] | Uganda | Study aim: To explore Ugandan mental health care worker’s perspectives and experiences on the sexual and reproductive health of people living with mental illness  Population: staff in psychiatric hospital | People with SMI have normal human sexual desires but staff sometimes focused on the abnormal aspects of this. Maintaining intimate relationships important but SMI seen as affecting sexual desire. Inhibited or excessive sexual desire commonly observed |
| Wainberg, 2007[61] | Brazil | Study aim: Characterize individual, institutional, and interpersonal factors that may affect HIV risk behavior in this population.  Population: people with SMI and staff in psychiatric hospital | SMI patients have sexual desire and deal with stigma |
| Yang 2023[62] | China | Study aim: Assess unmet sexual health needs of people with schizophrenia and the current level of support they receive from the health system in order to improve future support and tailor personal care plans.  Population: people with schizophrenia | The sexual needs of people with schizophrenia are often neglected, the environment doesn't provide sufficient private space for sexual activity and psychotic symptoms and side effects impair sexual function. Psychotic symptoms: Involuntary sexual abstinence and effect of symptoms on maintaining a relationship. Side effects of antipsychotics: Impaired sexual function, including low libido and difficulty reaching orgasm. Sex was considered an effective way to regulate emotion and to avoid negative emotions in daily life. Urgent hospital admission and frequent readmissions affect ability to maintain connection and increases feelings of isolation. Sex considered vital for maintaining relationships. Good sexual relationships were described as increasing happiness. Professionals described being unclear of the impact of schizophrenia on sexual life but felt the topic was too taboo to discuss with colleagues |

### Table 3.1 Contraception Use & Family Planning: Quantitative Studies

| Author, year | Country | Parameter | Contraception Use | |
| --- | --- | --- | --- | --- |
|  |  |  | **Women with SMI** | **Comparison** |
| Bursalioglu, 2013[63] | Turkey | Number not using contraception | 12 (35%) women with schizophrenia  8 (44%) women with BD | 3 (11%) women with depression  6 (17%) women without mental illness |
| Correa, 2020[64] | Colombia | Number feeling well informed about contraception | 32 (42.6%) women with BD  2 (50%) women with Schizophrenia | 14 (42.4%) men with BD  9 (18.7%) men with Schizophrenia |
| Desai, 2009[65] | India |  | Contraceptive issues were discussed with 15% of 135 women referred to a perinatal psychiatric clinic | No comparison |
| Eroglu, 2020 [66] |  | Number using any kind of contraception method  Number using traditional contraception methods | Significantly higher rates of contraception use in SMI group (<0.0011  n=56 (96.6%) women with BD  n=23 (40%) women with BD | n=45 (76.3%) women without mental illness  n=9 (15%) women without mental illness |
| Grover, 2019[67] | India | Number not using contraception | 26.2% of women | 35.7% of men |
| Guimarães, 2010[20] | Brazil | Number having unprotected sex at least once in their lifetime | 83.6% of women | 76.9% of men |
| Magalhães, 2009[68] | Brazil | Number using contraception | 58.8% of women with BD | No comparison |
| Marengo, 2015 (a)[69] | Argentina | Number using any effective contraceptive method | 90.7% of women with BD | 91.8% of women without BD |
| Marengo, 2015 (b) [22] | Argentina | Number with consistent condom use | 42.9% of women with BD | 40.4% of women without BD |
| Ozcan, 2014[70] | Turkey |  | 115 (47%) women with SMI they had used a contraceptive method in their last sexual intercourse.  51 (54%) women with SMI used coitus interruptus, 47 (23.4%) used intrauterine devices and 30 (14.9%) used condoms | No comparison |
| Pehlivanoglu, 2007 [71] | Turkey | Number reporting use of contraception during the last intercourse  Coitus interruptus was the most common method in all groups, followed by IUD, pill, and condom. Parenteral contraceptives were not known adequately, especially in patient groups. Calendar method, foam-cream-gel and diaphragm were among the least known methods. | 60.5% of patients with BD  68.6% of patients with schizophrenia | 75.5% of patients with depression  81.4% of women without mental illness |
| Peixoto, 2014[25] | Brazil | Number experiencing partner refusing condom,  Number always using a condom | 40.0% of women  11.2% of women | 23.6% of men  22.8% of men |
| Wainberg, 2008[27] | Brazil |  | 60.5% reported not using a condom due to their partners' preference. Other common reasons among women were: condoms unavailable at the time of intercourse (31.3%), trust in their partner(s) (25.0%), not being in the habit of using condoms (18.8%), and participant's own preference not to use condoms (18.8%). | No comparison |
| Zerihun, 2020[72] | Ethiopia |  | 56.6% of women with SMI reported that they had ever used contraception with 38.4% currently using at least one method of contraception. Of the women not currently using contraception, 60% had no intention to use contraception. | No comparison |
| Author, year | **Country** | **Parameter** | **Family Planning** | |
|  |  |  | **Women with SMI** | **Comparison** |
| Ceylan, 2019[73] | Turkey |  | 23.5% of healthcare professionals give family planning advice to people with schizophrenia. 18.2% said that individuals should decide for themselves whether to get an abortion (58.5% no, 23% don't know)  21.4% said that forced abortion and sterilisation for people with schizophrenia is not necessary (39% it is necessary but I don't support culturally, 39.6% it is necessary and I support culturally) | No comparison |
| Correa, 2020[64] | Colombia | Number receiving advice from psychiatry about family planning | 12 (16%) women with BD  1 (25%) women with Schizophrenia | 5 (15.1%) men with BD  3 (6.2%) men with Schizophrenia |
| Ozcan, 2014[70] | Turkey |  | 42.8% of the participants reported that their source of knowledge regarding contraceptive methods was neighbours and friends, and only 21.9% mentioned a health professional in this context. | No comparison |
| Pehlivanoglu, 2007[71] | Turkey |  | The patients in the schizophrenia group had less information on all contraceptive methods.  Discussing contraception and family planning with partner was less frequent in schizophrenics (60% no; 40% yes) and bipolar patients (50% no; 50% yes) | Discussing contraception and family planning with partner was more frequent in depression and control groups (60%, 90%, respectively). |
| Tunde, 2013[75] | Nigeria | Study aim: To identify the current status of knowledge and pattern of use of modern contraceptives in a clinic population.  Population: people with SMI | 88% of the women had good knowledge of contraception, 61% were interested in its use but 51% of them had not used any method and current use was just 27%. - Method of family planning known in decreasing order was: Male condom (68%); injectables (64%); the pills (56%); IUD (37%); and sterilization (16%). Less than half (48%) discussed family planning issues with their spouses, only 5% had ever received family planning information from the study clinic or hospital even though 81% desired to have such information provided by clinics. | No comparison |
| Zerihun, 2020[72] | Ethiopia |  | 68% of study participants had ever heard about contraception. Three most commonly known methods were the oral contraceptive pill (29.6%) the injectable (depot contraceptive) (29%), condoms (22.8%). Most frequently mentioned source of contraception information was health professionals (52.6%), a friend or neighbours (20.2%), school (10.5%) and the media (5.2%). | No comparison |

### Table 3.2 Contraception Use and Family Planning: Qualitative Studies

| Author, year | Country | Contraception use | |
| --- | --- | --- | --- |
|  |  | **Study Aim & Population** | **Qualitative Findings: Women with SMI** |
| Collins, 2001[28] | South Africa | Study aim: This paper presents data from a qualitative study of South African mental health care providers, examining their perceptions of the HIV risk and experiences of sexuality among women with SMI.  Population: Men and women who had direct contact and clinical or rehabilitation responsibilities for individuals with mental illness or administrative duties in provincial mental health care departments. | Routine prescription of Depo-Provera (contraceptive injection) for inpatients, condoms not always stocked. All providers believed that women with SMI should have access to contraception. Previously, contraception was given without consent. Now consent is taken but can be taken from the family if the patient refuses. There was a belief among providers that making condoms available would be interpreted as acknowledging and condoning the sexual activity. Many providers were enthusiastic about the use of abortion as an acceptable method of birth control if contraception failed or had not been used, and if the woman requested abortion. Sterilization was considered an important alternative to contraception for women with severe mental illnesses by some providers. |
| Pinto, 2007 [29] | Brazil | Study aim: to Investigate the beliefs, values, attitudes, and motivations related to behaviours that would provide basis for developing interventions for prevention of HIV among male and female mental health service users  Population: People with SMI and admin and patient facing staff | Staff reported instances of patients having sexual intercourse without condoms. |
| Sethuraman, 2019 [74] | India | Study aim: To explore knowledge, attitude and practice of contraception among women with schizophrenia.  Population: people with schizophrenia | 88.5% of women with schizophrenia had knowledge of any form of contraception and 51% had knowledge of more than one method of contraception (median 2, range 0-5). Female sterilization, intrauterine devices, condoms, and oral contraceptive pills were the most commonly known contraceptive methods.  74/96 (77.1%) had used contraception at some point in time. Sterilisation and intrauterine devices were the most commonly used methods (Female sterilisation 58.3%, intrauterine device 15.6%. No current users of contraception were using oral contraceptive pills, injectable, implants, female condom, or diaphragm. 22.9% had never used contraception |
| Shahvari, 2020[30] | Iran | Study aim: to explore factors which should be considered in designing a sexuality education package for severe mental illness.  Population: SMI patients, their family, clinical staff in a psychiatric unit and a general physician | Women not using condoms due to risk of being perceived negatively by partner. |
| Tumwakire, 2022[60] | Uganda | Study aim: To explore Ugandan mental health care worker’s perspectives and experiences on the sexual and reproductive health of people living with mental illness  Population: staff in psychiatric hospital | Reversible short term contraceptive methods (such as Depo injection and implant) were commonly given |
| Wainberg, 2007[61] | Brazil | Study aim: Characterize individual, institutional, and interpersonal factors that may affect HIV risk behaviour in this population.  Population: people with SMI and staff in psychiatric hospital | Participant experienced regret after sexual intercourse without condoms |
| Author, year | **Country** | **Family planning** | |
|  |  | **Study Aim & Population** | **Qualitative Findings: Women with SMI** |
| Bagadia, 2020[76] | India | Study aim: To determine factors that influence decision-making regarding pregnancy for women with SMI.  Population: people with schizophrenia | Lack of autonomy in making decisions about planning pregnancy and contraception |
| Nakigudde, 2013 [77] | Uganda | Study aim: To explore how family psychoeducation could be made culturally sensitive, feasible and appropriate for postpartum women with psychotic illness in Central Uganda.  Population: women with SMI, their caregivers and staff in psychiatric hospital | Inclusion of Family Planning education. Some caregivers and postpartum women believed that family planning education should be made part of the psychoeducation for postpartum women. |
| Sethuraman, 2019 [74] | India | Study aim: To explore knowledge, attitude and practice of contraception among women with schizophrenia.  Population: people with schizophrenia | 14.6% had unmet contraceptive needs with multiple reasons given for this including lack of awareness & not being given info. |
| Sibanyoni, 2022 [78] | South Africa | Study aim: To explore patient awareness of the teratogenic risk of sodium valproate  Population: people with BD | A small number of women had concerns about sodium valproate, that it was not their medication of choice and that taking it affected their family planning decisions. |
| Tumwakire, 2022[60] | Uganda | Study aim: To explore Ugandan mental health care worker’s perspectives and experiences on the sexual and reproductive health of people living with mental illness  Population: staff in psychiatric hospital | Due to the observed deranged sexuality, all participants noted to recommend contraception for all at risk patients. Some of the participants provided room for relatives and patients when stable to make contraception decisions while, some participants just decided for patients. Participants noted that being on contraception was in best interest for the patients and the community to prevent producing children who would not get appropriate care. |
| Zerihun, 2021 (a)[79] | Ethiopia | Study aim: Explore the family planning experiences and preferences and unmet needs of women with SMI who reside in a predominantly rural area of Ethiopia  Population: people with SMI | Reluctance to speak about family planning as most participants associated family planning with prevention of birth, rather than planned birth. Negative associations to condom use e.g. promiscuity. Inconsistent knowledge about contraception and barriers to timely access in primary care. Need for accessibility and privacy, and raised concerns about stigma, lack of adequate knowledge about family planning, and the need for special considerations in the family planning service. Most participants preferred to be provided with family planning services in a mental health clinic and by a mental health professional. The reason given was the need for the person advising on family planning to have adequate knowledge about mental health. A few suggested their home as another alternative service area for family planning in women with SMI and a preference for individual instead of group format. Unmet needs for awareness and access to for emergency family planning. |

Table 4.1 Sexual Violence: Quantitative Studies

| Author, year | Country | Parameter | Sexual violence | |
| --- | --- | --- | --- | --- |
|  |  |  | **Women with SMI** | **Comparison** |
| Afe, 2016[80] | Nigeria | Number reporting sexual assault | 19/77 (25%) of women | No comparison |
| Amr, 2012[81] | Egypt | Number with previous experience of sexual abuse | 3/37 (8.1%) of women | 16/61 (26.2%) of men |
| Chandra, 2003 (a) [16] | India | Number reporting having penetrative sex when they did not want to due to threat or force | 14% of women | No comparison |
| Chandra, 2003 (b) [82] | India | Number reporting at least one episode of coercive sex | 50/50 (100%) of women | No comparison |
| De Oliveira, 2012 [83] | Brazil | Number reporting experience of sexual violence in life | 26.6% of women | 12.5% of men |
| Incedere, 2017 [47] | Turkey | Number not facing sexual abuse | 75/116 (65.7%) of women | 72/84 (85.7%) of men |
| Lundberg, 2015[21] | Uganda | Number experiencing sexual violence by partner,  Number experiencing sexual violence by non partner | 37/153 (24.2%) of women 29/277 (10.5%) of women | 312/1497 (20.8%) of women without SMI  19/1774 (1.1%) of women without SMI |
| Mamabolo, 2012[84] | South Africa | Likelihood of being forced into sexual intercourse | Women significantly more likely | Men significantly less likely |
| Tomruk, 2006[26] | Turkey | Number experiencing forced intercourse | 10% of women | No comparison |
| Ozcan, 2014[70] | Turkey | Number experiencing forced intercourse by non-partner,  Number experiencing forced intercourse by partner | 11.3% of women  20.5% of women | No comparison |
| Peixoto, 2014[25] | Brazil | Number experiencing lifetime sexual violence | 28.8% of women | 12.4% of men |
| Zerihun, 2021 (b)[85] | Ethiopia |  | 25.6% of women reporting experience of physical violence also reported experiencing sexual violence | No comparison |

### Table 4.2. Sexual Violence: Qualitative Studies

| Author, year | Country | Sexual Violence | |
| --- | --- | --- | --- |
|  |  | **Study Aim & Population** | **Qualitative Findings: Women with SMI** |
| Abekah-Carter, 2022[86] | Ghana | Study aim: (a) to find out the impacts of the presence of persons with mental illness on the streets and (b) to ascertain the reasons accounting for homelessness among persons with mental illness.  Population: community members of homeless people | Reports of rape in homeless women with SMI and difficulty accessing medical care after rape. |
| Collins, 2006[87] | South Africa | Study aim: To examine those issues that challenge mental health care providers’ ability to intervene in the epidemic among their patients: their own views of psychiatric illness, the transitions occurring in the mental health care system, and shifting social attitudes toward sexuality.  Population: staff in psychiatric hospital | A challenge for providers was the perceived risk of sexual assault for people with mental illness in inpatient units due to their right to associate with the opposite sex. |
| Collins, 2001[28] | South Africa | Study aim: This paper presents data from a qualitative study of South African mental health care providers, examining their perceptions of the HIV risk and experiences of sexuality among women with SMI.  Population: Men and women who had direct contact and clinical or rehabilitation responsibilities for individuals with mental illness or administrative duties in provincial mental health care departments. | Reports of victimisation and rapes from women with SMI. |
| Ghebrehiwet, 2019[88] | Ethiopia | Study aim: By examining community perspectives on cultural explanatory models and experiences of SMI in Butajira, we seek to gain a better understanding of possible interventions aimed at improving case detection for women with mental illness in similar settings.  Population: people with SMI, their caregivers and community members | People who care for women with SMI noted they are especially vulnerable to rape. |
| Hall, 2019[89] | Timor-Leste | Study aim: to investigate the experiences of and opinions about social inclusion and exclusion of Timorese people with mental illness from the perspective of multiple stakeholders.  Population: (1) people with mental illness and their families; (2) mental health and social service providers; (3) government decision makers; (4) civil society members; and (5) other community members. | Sexual violence against women with mental illness noted. |
| Lundberg, 2012[57] | Uganda | Study aim: To understand how having a SMI may influence sexual risk behaviours and sexual health risks in a low-income sub-Saharan African country with high HIV prevalence.  Population: people with SMI | Five female participants described experiences of rape by non-partners after the first onset of their illness. At least three of these women were raped during illness episodes. |
| Poreddi, 2021[90] | India | Study aim: To explore women’s experiences of violence and their opinion on routine screening for domestic violence.  Population: people with SMI | Reports by women with SMI of incestuous sexual assault and violence precipitating psychiatric admissions and this resuming on discharge home or preventing them from wanting to return to family homes. |
| Tumwakire, 2022[60] | Uganda | Study aim: To explore Ugandan mental health care worker’s perspectives and experiences on the sexual and reproductive health of people living with mental illness.  Population: staff in psychiatric hospital | Mental health care workers expressed the concern that SMI increases sexual vulnerability with the majority of the female patients getting raped, including whilst admitted as inpatients. |
| Zerihun, 2021 (a)[79] | Ethiopia | Study aim: Explore the family planning experiences and preferences and unmet needs of women with SMI who reside in a predominantly rural area of Ethiopia  Population: people with SMI | Women faced vulnerabilities to sexual violence which led to unintended pregnancies. |

### Table 5.1. STI (non-HIV) Prevalence: Quantitative Studies

| Author, year | Country | Parameter | STI prevalence | |
| --- | --- | --- | --- | --- |
|  |  |  | **Women with SMI** | **Comparison** |
| Abayomi, 2013[15] | Nigeria | Number with previous STI diagnosis | 3/28 (11%) women | 8/74 (11%) men |
| Carey, 2007 [2] | India | Diagnosed with chlamydia,  Syphilis,  Hepatitis B,  Any STI | 31/287 (10.8%) women  7/375 (1.9%) women  6/375 (1.6%) women  50/375 (13.3%) women | 42/440 (9.6%) men  24/573 (4.2%) men  22/573 (3.8%) men  88/573 (15.4%) men |
| Carmo, 2013[91] | Brazil | Number with hepatitis C | 1.7% women | 3.4% men |
| Carmo, 2014 [92] | Brazil | Number with HBsAg seropositivity | 1.13% women | 3.02% men |
| De Oliveira, 2012[83] | Brazil | Number reporting lifetime STD diagnosis | 20.4% women | 26.4% men |
| Dutra, 2014[93] | Brazil | Number reporting previous STI diagnosis | 22.2% women | 29.8% men |
| Marengo, 2015 (b)[22] | Argentina | Number with previous STI diagnosis | 41.7% women | 33.2% women without SMI |
| Mpango, 2022 [11] | Uganda | OR for syphilis | 1.28 (CI 0.73; 2.25) women | 1 men |
| Zhang, 2018 (a)[94] | China |  | 11/29 (38%) patients with syphilis were female | 18/29 (62%) patients with syphilis were male |

### Table 5.2. STI (non-HIV) Prevalence: Qualitative Studies

| Author, year | Country | STI prevalence | |
| --- | --- | --- | --- |
|  |  | **Study Aim & Population** | **Qualitative Findings: Women with SMI** |
| Collins, 2001[28] | South Africa | Study aim: This paper presents data from a qualitative study of South African mental health care providers, examining their perceptions of the HIV risk and experiences of sexuality among women with SMI.  Population: Men and women who had direct contact and clinical or rehabilitation responsibilities for individuals with mental illness or administrative duties in provincial mental health care departments. | High rates of syphilis in hospital SMI population |
| Pinto, 2007[29] | Brazil | Study aim: to Investigate the beliefs, values, attitudes, and motivations related to behaviours that would provide basis for developing interventions for prevention of HIV among male and female mental health service users  Population: People with SMI and admin and patient facing staff | Staff observed SMI patients to be at higher risk of STI than general population |

### Table 6.1. Fertility, Pregnancy and Post-partum: Quantitative Studies

| Author, year | Country | Fertility | |
| --- | --- | --- | --- |
|  |  | **Women with SMI** | **Comparison** |
| Bhatia, 2004[95] | India | 67/89 (75.3%) of Indian women were childless | 73/101 (72.3%) Indian men were childless |
| Grover, 2019[67] | India | Mean number of conceptions was 2.41 in women | Mean number of conceptions 2.40 in men |
| Marengo, 2015 (a) [69] | Argentina | 52.4% of women with BD had ever been pregnant | 58.7% of women without BD had ever been pregnant |
| Terzian, 2006[96] | Brazil | The fertility rate for the women with SMI was 31.6% in the 25-44 age group and 28.6% in the 45+ age group. The fecundity of the general female population was 1.69. | The fertility rate for the general female population was 73.1% in the 25-44 age group and 87.3% in the 45+ age group. The fecundity of the general female population was 2.71. |
| Study | **Country** | **Pregnancy** | |
|  |  | **Women with SMI** | **Comparison** |
| Desai, 2009[65] | India | 44 (32%) of women with SMI had a history of accidental exposure to psychotropic drugs. Case of pregnant women not engaging in decision making around her pregnancy due to experiencing negative symptoms. Case of pregnant women afraid to take antipsychotics during pregnancy for fear of harm to foetus | No comparison |
| Eroglu, 2020 [66] | Turkey | No significant difference between groups in regularity of menstruation, number of times being pregnant and number of lost or interrupted pregnancies.  Total unplanned pregnancies in the women with bipolar disorder group was 49.52% | Women without bipolar disorder    Total unplanned pregnancies in the control group, it was 15.04% (no significance given). |
| Ozcan 2014[70] | Turkey | 80.7% of women experienced planned pregnancy and 51.4% experienced unplanned pregnancy | No comparison |
| Ozcan, 2018[97] | Turkey | Women with SMI were less likely to have experienced antenatal care during pregnancy and had a greater frequency of caesarean birth and were subject to more trauma. | Women without SMI were more likely to experience antenatal care and had a lower frequency of caesarean birth and trauma. |
| Correa, 2020 [64] | Colombia | Average age for the first pregnancy was 22 among women with BD and 32 among women with schizophrenia. | No comparison |
| Zerihun, 2020[72] | Ethiopia | 66.1% had a history of pregnancy. Of these women, 87.8% had experienced an unintended pregnancy. | No comparison |
| Author, year | **Country** | **The Postpartum Period** | |
|  |  | **Women with SMI** | **Comparison** |
| Ozcan, 2018[97] | Turkey | Postpartum mothers diagnosed with a mental health disorder were less likely to breastfeed, expressed more concerns about infant care, were subjected to more trauma during the postpartum period and relied on others to care for their babies more. | Postpartum mothers without mental illness were more likely to breastfeed their babies, expressed less concerns about infant care, were subject to less trauma and were more able to care for their babies themselves. |

### Table 6.2. Fertility, Pregnancy and Post-partum: Qualitative Studies

| Author, year | Country | Fertility | |
| --- | --- | --- | --- |
|  |  | **Study Aim & Population** | **Qualitative Findings** |
| Yu, 2022[98] | China | Study aim: To explore the experience of reproductive concerns through the perspective of women with schizophrenia.  Population: people with schizophrenia | Despite several difficulties faced in the fertility process, people still want to have children. Most of the participants considered parenting a rewarding process. Being a mother gave participants with schizophrenia more motivation to recover. |
| Study | **Country** | **Pregnancy** | |
|  |  | **Study Aim & Population** | **Qualitative Findings** |
| Aneja, 2020[99] | India | Study aim: To highlight the various ethical dilemmas that confront a psychiatrist while managing a patient from this group [pregnant women with SMI].  Population: a 25 year old pregnant women with schizophrenia and epilepsy | Dilemmas related to capacity to make decisions around keeping a pregnancy whilst psychotic and managing psychiatric medications during pregnancy |
| Bagadia, 2020[76] | India | Study aim: To determine factors that influence decision-making regarding pregnancy for women with SMI.  Population: people with schizophrenia | Decreased risk perception related to mental illness in the perinatal period and excessive focus on medication-related risk to the foetus. |
| Loganathan, 2022[100] | India | Study aim: To look at the disadvantages created by stigma that exist among Indian men and women who have schizophrenia.  Population: people with schizophrenia | Being coerced not to have children or being forced to abort for fear the child will also suffer from mental illness. |
| Poreddi, 2021[90] | India | Study aim: To explore women’s experiences of violence andtheir opinion on routine screening for domestic violence  Population: people with SMI | Results of pregnancies lost due to domestic violence. |
| Rezaie, 2020[59] | Iran | Study aim: to determine the unique post-discharge needs of Iranian women diagnosed with severe mental illness.  Population: people with SMI | Women wanted more education about pregnancy and the risk of medication on the developing foetus |
| Sibanyoni, 2022[78] | South Africa | Study aim: To explore patient awareness of the teratogenic risk of sodium valproate  Population: people with BD | 20/23 (87%) knew they needed to follow steps should they be planning a pregnancy or if they fall pregnant whilst on Epilim but fewer understood the potential teratogenic effects 13/23 (57%). |
| Yu, 2022[98] | China | Study aim: To explore the experience of reproductive concerns through the perspective of women with schizophrenia.  Population: people with schizophrenia | People with schizophrenia may have to manage psychiatric symptoms by taking medication during pregnancy. However, medications can be hazardous and most participants expressed fear that they may affect their unborn child. |
| Zerihun, 2021 (a)[79] | Ethiopia | Study aim: Explore the family planning experiences and preferences and unmet needs of women with SMI who reside in a predominantly rural area of Ethiopia  Population: people with SMI | Fear of relapse during or after birth, fears about ability to parent and effect of medication on babies. 11/16 (69%) participants felt women with SMI should not give birth. Fears around being able to care for a child. 1/16 (6%) feared a baby's health could be affected by medication but had not raised these concerns with health professionals. Concerns that community members felt women with SMI should not have children. |
| Author, year | **Country** | **The Postpartum Period** | |
|  |  | **Study Aim & Population** | **Qualitative Findings: Women with SMI** |
| Nakigudde, 2013 [77] | Uganda | Study aim: To explore how family psychoeducation could be made culturally sensitive, feasible and appropriate for postpartum women with psychotic illness in Central Uganda.  Population: women with SMI, their caregivers and staff in psychiatric hospital | Women with SMI being told not to breastfeed as SMI could be passed onto child through breastmilk. |
| Yu, 2022[98] | China | Study aim: To explore the experience of reproductive concerns through the perspective of women with schizophrenia.  Population: people with schizophrenia | During breastfeeding, changes in hormone levels can make the mother's body more vulnerable. Additionally, breastfeeding prevents mothers from having a regular routine and getting adequate sleep, which results in health risks. |

1. Data collected using a bespoke questionnaire rather than a validated tool
